# The pervasive association between political ideology and COVID-19 vaccine uptake in Brazil: an ecologic study

**DOI:** 10.1101/2022.10.24.22281482

**Authors:** Gabriel J. Seara-Morais, Thiago J. Avelino-Silva, Marcia Couto, Vivian I. Avelino-Silva

## Abstract

**Background:** Studies suggest vaccine hesitancy is an increasingly significant phenomenon in Brazil and other countries. Moreover, political ideologies have emerged as an influencing factor for vaccine hesitancy during the COVID-19 pandemic.

**Methods:** In this study, we use information from publicly available databases to investigate the association between political alignment, depicted by the percentage of Bolsonaro voters in the presidential elections of 2018 and 2022, and COVID-19 vaccination in Brazilian municipalities, adjusted for human development index (HDI) scores and basic sociodemographic characteristics of voters.

**Findings:** For both the 2018 and 2022 elections, higher percentages of Bolsonaro voters were significantly associated with a lower vaccination index after adjustment for voters’ sociodemographic characteristics. We also found a statistically significant interaction between the percentage of Bolsonaro voters and HDI, with a more significantly detrimental effect of the right-wing political stance in municipalities in the lower HDI quartile.

**Interpretation:** Our study highlights what may be the beginning of a new scenario with unforeseen challenges for vaccine programs: the politicization of vaccines. Strategies to face these challenges should include joint efforts from governments and civil society for a common public health goal.

**Funding:** This manuscript received no specific funding

## Introduction

Mass vaccination has had a crucial impact on worldwide public health in the past decades. In Brazil, the establishment of the National Immunization Program (Programa Nacional de Imunizações, PNI) in 1975 grounded the implementation of vaccines as an official and state-supported health policy.^1^ As a result of coordinated efforts to expand access to vaccines, the mean vaccination coverage among Brazilian children younger than one year increased from 50% before the program to more than 90% by the late 1990s. Concurrently, a sharp reduction in cases and deaths due to vaccine-preventable diseases was registered in the country.^2–4^

Despite the unequivocal benefits of vaccination, vaccine coverage has been falling in Brazil in the past few years, particularly since 2015.^5–7^ Non-adherence to vaccination recommendations can result from access barriers, including issues related to patient mobility and transportation, costs, working hours in vaccination clinics, shortage of supplies, and unawareness about recommended vaccines in distinct situations. However, in some cases, vaccines are voluntarily avoided or postponed after deliberate assessment and decision by the patient (or by a parent or legal guardian). This scenario has been referred to as “vaccine hesitancy” and has been observed in several developed countries in Europe, the USA, Canada, Japan, and Australia.^8–10^ Some studies suggest that vaccine hesitancy is an increasingly significant phenomenon in Brazil, especially in subgroups with higher income and education.^5,7,11–14^

More recently, during the COVID-19 pandemic, political views have emerged as an additional influencing factor for vaccine hesitancy. For example, a study in the USA showed that counties with a higher percentage of votes for the Republican party had lower vaccination coverage and higher rates of COVID-19 cases and deaths.^15^ In Brazil, president Jair Bolsonaro refused to receive the COVID-19 vaccine and declared he would not give the vaccine to his daughter; moreover, he spread misinformation and conspiracy theories about the vaccine and made several declarations opposing recommendations from official health organizations during the pandemic.^16–18^ These attitudes may have influenced overall adherence to the COVID-19 vaccination campaign in Brazil, particularly among Bolsonaro’s political supporters. In agreement with this hypothesis, two recent studies showed that Brazilian municipalities supporting Jair Bolsonaro in the 2018 elections were less compliant with social distancing measures in the first pandemic wave and had higher COVID-19 mortality rates, particularly during the second wave of the disease in 2021.^19,20^ It is also plausible to assume that the detrimental influence of political ideology on vaccine acceptance and uptake might be heterogenous across different municipalities based on sociodemographic characteristics.

In this study, we used information from publicly available databases to investigate the association between political alignment, depicted by the percentage of Bolsonaro voters in the presidential elections of 2018 and 2022, and COVID-19 vaccination in Brazilian municipalities, adjusted for human development index (HDI) and basic demographic characteristics of voters.

## Methods

In this cross-sectional, ecologic study, the primary predictor of interest was the percentage of votes for Jair Bolsonaro in the first round of the 2018 and 2022 elections;^21^ the primary endpoint was the COVID-19 vaccination index, calculated as the number of COVID-19 vaccine doses administered up to September 2022^22^ divided by the number of inhabitants in each municipality according to estimates from July 2021.^23^ We also obtained municipal-level data on the HDI,^24^ categorized into quartiles, with the highest quartile corresponding to municipalities with higher socioeconomic development, the percentage of male voters, the percentage of voters who were older than 50 years, and the percentage of voters with middle school education or less, using publicly available, de-identified databases.^21^

Characteristics of Brazilian municipalities were described using counts, percentages, medians, and interquartile ranges (IQR), overall and according to COVID-19 vaccination index quartiles. The association between the percentage of Bolsonaro voters and the vaccination index was investigated using linear regression models with robust standard errors, adjusted for HDI, the percentage of male voters, the percentage of voters who were older than 50 years old, and the percentage of voters with middle school education or less. In addition, we explored whether the effect of the percentage of Bolsonaro voters on the COVID-19 vaccination index was modified in different quartiles of HDI using an interaction term. We used scatter plots and Pearson coefficients to investigate the correlation between the percentage of Bolsonaro voters and the vaccination index according to quartiles of HDI. We used Stata 15·1 (StataCorp. College Station, TX: StataCorp LP) for all analyses, with a 0·05 significance level.

Per Resolution 510/2016 of the Brazilian National Health Council, our local Ethics Committee exempted our study from obtaining informed consent since we used exclusively publicly available, de-identified information.

## Results

Out of 5,570 Brazilian municipalities, 5,563 were included in the analysis based on the availability of data on the total population and number of COVID-19 vaccine doses. Table 1 presents sociodemographic characteristics of the municipalities included in the analysis, overall and according to the COVID-19 vaccination index quartiles. More than 60% of all municipalities were located in the Northeast and Southeast regions of the country. The percentage of male voters was close to 50%, and the percentage of voters with middle school education or less was close to 53%, overall and in each of the vaccination index quartiles. The percentage of voters who were older than 50 years old increased with increasing vaccination index quartiles. HDI distribution varied across COVID-19 vaccination index quartiles, with more municipalities with higher HDI in the higher vaccination index quartiles. The overall percentage of Bolsonaro voters was 41% in 2018 and 2022, with a lower percentage of votes in municipalities in the lower quartile of the COVID-19 vaccination index.

**Table 1:** Municipal-level characteristics according to quartiles of COVID-19 vaccination index.

|  | All municipalities<br>N=5563 | Vaccination index<br>Quartile 1<br>N=1391 | Vaccination index<br>Quartile 2<br>N=1391 | Vaccination index<br>Quartile 3<br>N=1390 | Vaccination index<br>Quartile 4<br>N=1391 |
| --- | --- | --- | --- | --- | --- |
| Region (%) |  |  |  |  |  |
| North | 448 (8) | 366 (26) | 59 (4) | 17 (1) | 6 (<1) |
| Northeast | 1792 (32) | 541 (39) | 487 (35) | 433 (31) | 331 (24) |
| Central-west | 467 (8) | 177 (13) | 116 (8) | 80 (6) | 94 (7) |
| Southeast | 1666 (30) | 182 (13) | 403 (29) | 499 (36) | 582 (42) |
| South | 1190 (21) | 125 (9) | 326 (23) | 361 (26) | 378 (27) |
| Median population size (IQR) | 11741<br>(5453-25769) | 17584<br>(8965-34653) | 15327<br>(7493-31589) | 11343<br>(5467-24237) | 5447<br>(3228-11609) |
| Percentage of male voters (IQR) | 49 (48-51) | 50 (48-51) | 49 (48-50) | 49 (48-50) | 50 (49-51) |
| Percentage of voters older than 50 years old (IQR) | 38 (34-42) | 33 (30-37) | 38 (34-41) | 39 (35-49) | 42 (37-45) |
| Percentage of voters with ≤ middle school education (IQR) | 53 (46-59) | 55 (48-60) | 53 (46-59) | 52 (45-59) | 52 (46-58) |
| Human development index quartile (%) |  |  |  |  |  |
| First | 1397 (25) | 582 (42) | 332 (24) | 277 (20) | 206 (15) |
| Second | 1386 (25) | 427 (31) | 364 (26) | 293 (21) | 302 (22) |
| Third | 1412 (25) | 250 (18) | 342 (25) | 384 (28) | 436 (31) |
| Fourth | 1359 (25) | 131 (9) | 348 (25) | 434 (31) | 446 (32) |
| Percentage of Bolsonaro voters in 1<sup>st</sup> round 2018 (IQR) | 41 (21-55) | 32 (19-50) | 41 (21-56) | 44 (21-56) | 45 (25-55) |
| Percentage of Bolsonaro voters in 1<sup>st</sup> round 2022 (IQR) | 41 (24-53) | 35 (23-50) | 41 (24-54) | 42 (23-54) | 44 (26-54) |

IQR, interquartile range

Effect estimates obtained in multivariable models addressing the association between the percentage of Bolsonaro voters in 2018 and 2022 and the COVID-19 vaccination index adjusted for covariates and including an interaction term with HDI are presented in Table 2. For both elections, higher percentages of Bolsonaro voters were significantly associated with a lower vaccination index; moreover, the harmful effect of each percent increase in Bolsonaro voters was greater in the lowest quartile of HDI compared to the highest quartile in 2018; similarly, the harmful effect of each percent increase in Bolsonaro voters was greater in the first and second quartiles of HDI compared to the highest quartile in 2022. In 2018, each 1% increase in Bolsonaro voters was associated with a mean 0·11-unit reduction in the vaccination index for municipalities in the fourth HDI quartile and a mean 0·22-unit reduction in vaccination index for municipalities in the first HDI quartile (interaction p-value <0·001). In 2022, each 1% increase in Bolsonaro voters was associated with a mean 0·09-unit reduction in vaccination index for municipalities in the fourth HDI quartile, a mean 0·21-unit reduction in vaccination index for municipalities in the first HDI quartile (interaction p-value <0·001), and a mean 0·14-unit reduction in vaccination index for municipalities in the second HDI quartile (interaction p-value =0·001).

**Table 2:** Effect estimates in multivariable models for the COVID-19 vaccination index, including the interaction between the percentage of Bolsonaro voters and the human development index.

| Variables | 2018 election |  | 2022 election |  |
| --- | --- | --- | --- | --- |
|  | Coefficient<br>(95% CI) | p-value | Coefficient<br>(95% CI) | p-value |
| Change in mean vaccination index for each 1% increase of male voters | 0.39<br>(-0.44 to 1.21) | 0.357 | 1.20<br>(0.37 to 2.02) | 0.005 |
| Change in mean vaccination index for each 1% increase of voters with ≤ middle school education | -0.59<br>(-0.82 to -0.35) | <0.001 | -0.49<br>(-0.73 to -0.26) | <0.001 |
| Change in mean vaccination index for each 1% increase of voters older than 50 years old | 4.97 (4.66-5.28) | <0.001 | 4.78<br>(4.48 to 5.09) | <0.001 |
| Change in mean vaccination index for each 1% increase of Bolsonaro voters* | -0.11<br>(-0.13 to -0.08) | <0.001 | -0.09<br>(-0.11 to -0.07) | <0.001 |
| Change in mean vaccination index for each category of HDI# |  |  |  |  |
| First | -0.56 (-0.66 to -0.47) | <0.001 | -0.47 (-0.56 to -0.39) | <0.001 |
| Second | -0.28 (-0.34 to -0.21) | <0.001 | -0.24 (-0.31 to -0.18) | <0.001 |
| Third | -0.12 (-0.18 to -0.06) | <0.001 | -0.09 (-0.15 to -0.04) | 0.001 |
| Fourth | Reference | - | - | - |
| Interaction term: Mean difference in the effect of each 1% increase of Bolsonaro voters for each HDI category |  |  |  |  |
| First | -0.11 (-0.14 to -0.07) | <0.001 | -0.12 (-0.15 to -0.08) | <0.001 |
| Second | -0.02 (-0.05 to 0.01) | 0.216 | -0.05 (-0.08 to -0.02) | 0.001 |
| Third | 0.02 (-0.01 to 0.05) | 0.214 | 0.00 (-0.03 to 0.03) | 0.828 |
| Fourth | Reference | - | Reference | - |

HDI: human development index
\*Among municipalities in the fourth (higher) human development index quartile
#Among municipalities with mean percentage of Bolsonaro voters

The percentage of male voters had no statistically significant effect on the vaccination index in the model, including the 2018 election results; however, higher percentages of male voters were associated with a higher COVID-19 vaccination index in the model, including the 2022 election results. Higher percentages of voters with middle school education or less were significantly associated with a lower vaccination index in both models, whereas higher percentages of voters older than 50 years old were significantly associated with a higher vaccination index in both models. Finally, lower quartiles of HDI were significantly associated with lower vaccination index in both models.

The correlations between the percentage of Bolsonaro voters in 2018 and 2022 and the COVID-19 vaccination index in Brazilian municipalities, according to human development index quartiles, are presented in Figure 1. As observed in the multivariable models, the inverse correlation was stronger in municipalities in the lowest HDI quartile for both the 2018 and 2022 elections.

**Figure 1:**
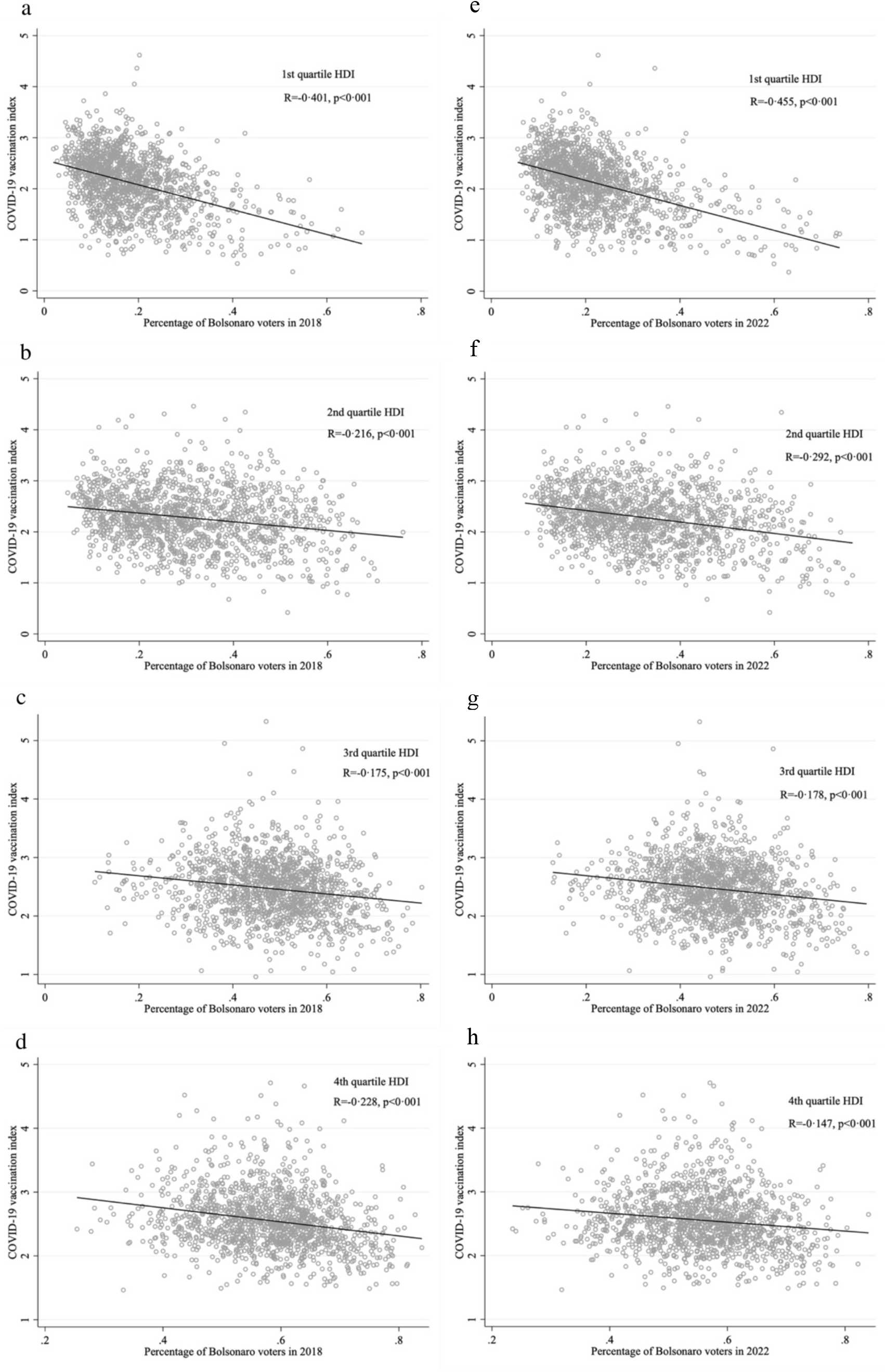
**Scatter plots depicting the correlation between the percentage of Bolsonaro voters in 2018 (panels a-d) and 2022 (panels e-h) and the COVID-19 vaccination index in Brazilian municipalities, according to human development index quartiles (plots including municipalities with vaccination index <6)**

## Discussion

In this cross-sectional ecologic study including Brazilian municipalities as units of observation, we showed that political ideology, depicted by the percentage of votes for the right-wing candidate Jair Bolsonaro, is significantly and inversely associated with COVID-19 vaccine uptake after adjusting for sociodemographic characteristics of the voters. We also showed a statistically significant interaction between the percentage of Bolsonaro voters and HDI scores, with a more significantly detrimental effect of the right-wing political stance in municipalities in the lower HDI quartile.

Our findings parallel results from a study completed in the USA, which demonstrated that counties with higher percentages of votes for the Republican party in the 2016 presidential election had lower COVID-19 vaccination coverage.^15^ Notably, Donald Trump and Jair Bolsonaro, both in line with a far-right wing denialist rhetoric,^25^ and respective heads of state of the United States and Brazil during the most challenging periods of the COVID-19 pandemic, expressed similar attitudes towards non-pharmacological prevention strategies and COVID-19 vaccines.^26^ For instance, both presidents opposed the adoption of facial masks and physical distancing and mobility restrictions;^27–31^ both repelled the implementation of vaccines produced in China;^32–34^ both manifested mistrust regarding COVID-19 vaccines;^35,36^ and both concealed information regarding their COVID-19 vaccination status.^37,38^ Our results are also supported by a recent survey study published by Paschoalloto et al., showing that willingness to be vaccinated for COVID-19 is strongly associated with political orientation.^39^ Our study provides additional evidence on the pervasive influence political ideologies can have on COVID-19 vaccination in Brazil and shows that municipalities in more vulnerable socioeconomic conditions seem even more susceptible to this effect. It is further disquieting that lower vaccination rates will likely lead to a higher disease burden in these municipalities, exacerbating prevailing social disparities.

While COVID-19 vaccine coverage is more directly relevant to the middle and long-term control of COVID-19,^40^ uptake of other vaccines has also been indirectly affected by the recent pandemic scenario. Several studies suggest that adherence to routine vaccines has dropped since the onset of the COVID-19 pandemic.^41–45^ Disruptions in healthcare services likely intensified barriers to vaccination in several settings. Nonetheless, it is also possible that skepticism towards COVID-19 vaccines built up distrust and hesitancy towards other vaccines, as suggested by a previous study on the influenza vaccine.^46^ Consequently, escalating challenges related to reductions in vaccine coverage, including outbreaks of vaccine-preventable diseases, might happen even after the re-establishment of routine care in vaccine clinics affected by the pandemic. Reluctant attitudes toward the COVID-19 vaccine may have reinforced the growing phenomenon of vaccine hesitancy.

Our study had limitations. First, we used HDI data from 2010 since more recent municipal-level information was unavailable from official sources. Second, age and schooling were analyzed as binary variables since more granularity or individual-level data could not be obtained from available datasets. Third, we used an ecologic design, which could be prone to ecologic fallacy and confounding. Even so, we were able to include data from most Brazilian municipalities and investigate interactions between the percentage of Bolsonaro voters and HDI on the COVID-19 vaccination index, adjusted for other sociodemographic covariates. To our knowledge, this is the first study addressing such an association in Brazil. Furthermore, we used data from both the 2018 and 2022 elections and found similar results, supporting that political views before and after the COVID-19 vaccine rollout were associated with our vaccination index.

There are several implications for our results. For over two decades, Brazil had been able to provide a robust public policy within the PNI, achieving high vaccination coverage for most vaccines and establishing a widespread culture of vaccination.^2,47^ However, in recent years the rates of vaccination have been dropping,^48^ with consequences that included a recent outbreak of measles in the state of Sao Paulo, in 2019.^49^ While barriers to vaccine access should still be confronted, vaccine hesitancy seems to be an increasingly concerning issue, enhanced by political ideologies and potentially affecting communities inequitably.^34,50^ Therefore, our study highlights what may be the beginning of a new scenario with unforeseen challenges for the PNI: the politicization of vaccines. Until recently, the Program recorded fairly high adherence to vaccines recommended in the national immunization calendar, and subsequent widespread reductions in coverage appeared unrelated to political affiliations or beliefs. However, in the current political scenario in Brazil, vaccines have shifted from ‘a health issue’ to ‘a political issue’. This could be the outcome of rhetorical disputes over the pandemic, which went beyond health and sanitation issues and strengthened the contemporary clash of worldviews on human relations, society’s organization, the role of governments, and the economy.^51,52^ Therefore, initiatives to address these difficulties should include collaborative efforts by governments and civil society toward a common goal that prioritizes public health regardless of individual political preferences.

## Data Availability

This study used publicly available, de-identified data only. Data sources are provided in the reference list

https://sig.tse.jus.br/ords/dwapr/seai/r/sig-eleicao-resultados/home?session=15363086146230

https://infoms.saude.gov.br/extensions/DEMAS_C19_Vacina_v2/DEMAS_C19_Vacina_v2.html

https://www.ibge.gov.br/estatisticas/sociais/populacao/9103-estimativas-de-populacao.html?=&t=resultados

http://www.atlasbrasil.org.br/ranking

## References

1 Silva Junior JB da. 40 anos do Programa Nacional de Imunizações: uma conquista da Saúde Pública brasileira. Epidemiol E Serviços Saúde 2013; 22: 7–8.

2 Hochman G. Vacinação, varíola e uma cultura da imunização no Brasil. Ciênc Saúde Coletiva 2011; 16: 375–86.

3 Domingues CMAS, Teixeira AM da S. Coberturas vacinais e doenças imunopreveníveis no Brasil no período 1982-2012: avanços e desafios do Programa Nacional de Imunizações. Epidemiol E Serviços Saúde 2013; 22: 9–27.

4 Temporão JG. O Programa Nacional de Imunizações (PNI): origens e desenvolvimento. História Ciênc Saúde-Manguinhos 2003; 10: 601–17.

5 Barata RB, Sampaio de Almeida Ribeiro MC, de Moraes JC, Flannery B, on behalf of the Vaccine Coverage Survey 2007 Group. Socioeconomic inequalities and vaccination coverage: results of an immunisation coverage survey in 27 Brazilian capitals, 2007–2008. J Epidemiol Community Health 2012; 66: 934–41.

6 de Moraes JC, de Almeida Ribeiro MCS, Simões O, de Castro PC, Barata RB. Qual é a cobertura vacinal real? Epidemiol E Serviços Saúde 2003; 12. DOI:10.5123/S1679-49742003000300005.

7 Silveira MF, Buffarini R, Bertoldi AD, et al. The emergence of vaccine hesitancy among upper-class Brazilians: Results from four birth cohorts, 1982–2015. Vaccine 2020; 38: 482–8.

8 MacDonald NE. Vaccine hesitancy: Definition, scope and determinants. Vaccine 2015; 33: 4161–4.

9 Larson HJ, Jarrett C, Eckersberger E, Smith DMD, Paterson P. Understanding vaccine hesitancy around vaccines and vaccination from a global perspective: A systematic review of published literature, 2007–2012. Vaccine 2014; 32: 2150–9.

10 Siddiqui M, Salmon DA, Omer SB. Epidemiology of vaccine hesitancy in the United States. Hum Vaccines Immunother 2013; 9: 2643–8.

11 Barbieri CLA, Couto MT. Decision-making on childhood vaccination by highly educated parents. Rev Saúde Pública 2015; 49. DOI:10.1590/S0034-8910.2015049005149.

12 Brown AL, Sperandio M, Turssi CP, et al. Vaccine confidence and hesitancy in Brazil. Cad Saúde Pública 2018; 34. DOI:10.1590/0102-311×00011618.

13 Gowda C, Dempsey AF. The rise (and fall?) of parental vaccine hesitancy. Hum Vaccines Immunother 2013; 9: 1755–62.

14 Sato APS. What is the importance of vaccine hesitancy in the drop of vaccination coverage in Brazil? Rev Saúde Pública 2018; 52: 96.

15 Albrecht D. Vaccination, politics and COVID-19 impacts. BMC Public Health 2022; 22: 96.

16 Daniels JP. Health experts slam Bolsonaro’s vaccine comments. The Lancet 2021; 397: 361.

17 Martins-Filho PR, Barberia LG. The unjustified and politicized battle against vaccination of children and adolescents in Brazil. Lancet Reg Health - Am 2022; 8: 100206.

18 Fonseca EM da, Nattrass N, Lazaro LLB, Bastos FI. Political discourse, denialism and leadership failure in Brazil’s response to COVID-19. Glob Public Health 2021; 16: 1251–66.

19 Xavier DR, Lima e Silva E, Lara FA, et al. Involvement of political and socio-economic factors in the spatial and temporal dynamics of COVID-19 outcomes in Brazil: A population-based study. Lancet Reg Health - Am 2022; 10: 100221.

20 Ajzenman N, Cavalcanti T, Da Mata D. More Than Words: Leaders’ Speech and Risky Behavior during a Pandemic. 2020; published online April 22. DOI:10.2139/ssrn.3582908.

21 SIG Eleição - Resultados. https://sig.tse.jus.br/ords/dwapr/seai/r/sig-eleicao-resultados/home?session=15363086146230 (accessed Oct 20, 2022).

22 Vacinometro COVID-19. https://infoms.saude.gov.br/extensions/DEMAS_C19_Vacina_v2/DEMAS_C19_Vacina_v2.html (accessed Oct 20, 2022).

23 Estimativas da população residente para os municípios e para as unidades da federação | IBGE. https://www.ibge.gov.br/estatisticas/sociais/populacao/9103-estimativas-de-populacao.html?=&t=resultados (accessed Oct 20, 2022).

24 Atlas Brasil. http://www.atlasbrasil.org.br/ranking (accessed Oct 20, 2022).

25 Hallal PC. SOS Brazil: science under attack. The Lancet 2021; 397: 373–4.

26 Barbara V. Opinion | Brazil Is Brilliant at Vaccinations. So What Went Wrong This Time? N. Y. Times. 2021; published online Feb 28. https://www.nytimes.com/2021/02/28/opinion/brazil-covid-vaccines.html (accessed Oct 20, 2022).

27 Coronavirus: Donald Trump vows not to order Americans to wear masks. BBC News. 2020; published online July 18. https://www.bbc.com/news/world-us-canada-53453468 (accessed Oct 20, 2022).

28 Murakawa F, Schuch M. ‘Aqui, é proibido máscara’, diz Bolsonaro a forrozeiros no Planalto. Valor Econômico. https://valor.globo.com/brasil/noticia/2021/12/13/aqui-e-proibido-mascara-diz-bolsonaro-a-forrozeiros-no-planalto.ghtml (accessed Oct 20, 2022).

29 Côrtes G, Caramuru P. Bolsonaro pede parecer para desobrigar uso de máscara a vacinados e quem já foi infectado - Saúde. O Estado Paulo. https://saude.estadao.com.br/noticias/geral,bolsonaro-diz-que-pediu-parecer-para-desobrigar-uso-de-mascara-a-vacinados-e-quem-ja-foi-infectado,70003743354 (accessed Oct 20, 2022).

30 Bolsonaro culpa distanciamento social pela inflação, ironiza Covid e diz que herdou ‘casa desarrumada’. Carta Cap. 2021; published online Sept 13. https://www.cartacapital.com.br/cartaexpressa/bolsonaro-culpa-distanciamento-social-pela-inflacao-ironiza-covid-e-diz-que-herdou-casa-desarrumada/ (accessed Oct 20, 2022).

31 Trump continues to flout social distancing guidelines even as he urges others to follow them. Wash. Post. https://www.washingtonpost.com/politics/trump-social-distancing/2020/08/06/5df4998c-d746-11ea-9c3b-dfc394c03988_story.html (accessed Oct 20, 2022).

32 Gullino D. Veja 10 vezes em que Bolsonaro criticou a CoronaVac. O Globo. 2021; published online Jan 18. https://oglobo.globo.com/politica/veja-10-vezes-em-que-bolsonaro-criticou-coronavac-24843568 (accessed Oct 20, 2022).

33 A Chinese Vaccine Could Save American Lives. Bloomberg.com. 2020; published online Oct 25. https://www.bloomberg.com/opinion/articles/2020-10-25/chinese-vaccine-for-covid-19-trump-should-agree-to-test-in-u-s (accessed Oct 20, 2022).

34 Gramacho WG, Turgeon M. When politics collides with public health: COVID-19 vaccine country of origin and vaccination acceptance in Brazil. Vaccine 2021; 39: 2608–12.

35 Bolsonaro sobre vacina de Pfizer: ‘Se você virar um jacaré, é problema de você’ -18/12/2020 - UOL Notícias. https://noticias.uol.com.br/ultimas-noticias/afp/2020/12/18/bolsonaro-sobre-vacina-de-pfizer-se-voce-virar-um-jacare-e-problema-de-voce.htm (accessed Oct 20, 2022).

36 Analysis | Trump follows his base toward rationalized vaccine skepticism. Wash. Post. https://www.washingtonpost.com/politics/2021/07/19/trump-follows-his-base-toward-rationalized-vaccine-skepticism/ (accessed Oct 20, 2022).

37 Haberman M. Trump and his wife received coronavirus vaccine before leaving the White House. N. Y. Times. 2021; published online March 1. https://www.nytimes.com/2021/03/01/us/politics/donald-trump-melania-coronavirus-vaccine.html (accessed Oct 20, 2022).

38 Martins L, de Andrade H. Mesmo dizendo que liberou acesso, Bolsonaro mantém sigilo de sua vacinação. https://noticias.uol.com.br/politica/ultimas-noticias/2022/10/19/jair-bolsonaro-sigilo-cartao-vacinacao.htm (accessed Oct 20, 2022).

39 Paschoalotto MAC, Costa EPPA, Almeida SV de, et al. Running away from the jab: factors associated with COVID-19 vaccine hesitancy in Brazil. Rev Saúde Pública 2021; 55: 97.

40 Moghadas SM, Vilches TN, Zhang K, et al. The Impact of Vaccination on Coronavirus Disease 2019 (COVID-19) Outbreaks in the United States. Clin Infect Dis 2021; 73: 2257–64.

41 Matos CC de SA, Barbieri CLA, Couto MT. Covid-19 and its impact on immunization programs: reflections from Brazil. Rev Saúde Pública 2020; 54: 114.

42 Guglielmi G. Pandemic drives largest drop in childhood vaccinations in 30 years. Nature 2022; 608: 253–253.

43 COVID-19 pandemic fuels largest continued backslide in vaccinations in three decades - PAHO/WHO | Pan American Health Organization. https://www.paho.org/en/news/15-7-2022-covid-19-pandemic-fuels-largest-continued-backslide-vaccinations-three-decades (accessed Oct 20, 2022).

44 SeyedAlinaghi S, Karimi A, Mojdeganlou H, et al. Impact of COVID – 19 pandemic on routine vaccination coverage of children and adolescents: A systematic review. Health Sci Rep 2022; 5. DOI:10.1002/hsr2.516.

45 Silveira MM, Conrad NL, Leivas Leite FP. Effect of COVID-19 on vaccination coverage in Brazil. J Med Microbiol 2021; 70. DOI:10.1099/jmm.0.001466.

46 Leuchter RK, Jackson NJ, Mafi JN, Sarkisian CA. Association between Covid-19 Vaccination and Influenza Vaccination Rates. N Engl J Med 2022; 386: 2531–2.

47 Domingues CMAS, Maranhão AGK, Teixeira AM, Fantinato FFS, Domingues RAS. 46 anos do Programa Nacional de Imunizações: uma história repleta de conquistas e desafios a serem superados. Cad Saúde Pública 2020; 36: e00222919.

48 Césare N, Mota TF, Lopes FFL, et al. Longitudinal profiling of the vaccination coverage in Brazil reveals a recent change in the patterns hallmarked by differential reduction across regions. Int J Infect Dis 2020; 98: 275–80.

49 Makarenko C, San Pedro A, Paiva NS, Santos JPC dos, Medronho R de A, Gibson G. Ressurgimento do sarampo no Brasil: análise da epidemia de 2019 no estado de São Paulo. Rev Saúde Pública 2022; 56: 50.

50 Fernandez M, Matta G, Paiva E. COVID-19, vaccine hesitancy and child vaccination: Challenges from Brazil. Lancet Reg Health - Am 2022; 8: 100246.

51 Bosco E, Igreja RL, Valladares L, editors. A América Latina frente ao Governo da COVID-19: desigualdades, crises, resistência, 1st edn. Brasília, DF, 2022 https://landportal.org/node/102355.

52 Romano JO, Bittencourt TP, Balthazar PAA, et al. A pandemia da Covid-19 como acontecimento e a disputa de discursos. Monde Dipl. Bras. https://diplomatique.org.br/o-virus-nao-e-democratico-a-pandemia-da-covid-19-como-acontecimento-e-a-disputa-de-discursos/ (accessed Oct 20, 2022).

